# Population-based urinary pesticide biomonitoring in rural Wisconsin: Longitudinal patterns and determinants of glyphosate, AMPA, and 2,4-D, and other modern use pesticides

**DOI:** 10.64898/2026.08.26.26361410

**Authors:** Amy A. Schultz, Meshel Lange, Brandon Shelton, Erin Meinholz, Doug Esselman, Edward Paulsen, Amanda Haban, Veronica Kesner, Marcy Rowe, Richard Burke, Christopher Tisler, Carrie Tomasallo

## Abstract

**Background:** Population-based biomonitoring of contemporary-use pesticides remains limited in the United States, particularly in rural agricultural regions, and few studies have repeated measurements within the same individuals over time.

**Methods:** We analyzed 28 urinary pesticide-related biomarkers among 600 adults from the population-based Survey of the Health of Wisconsin with archived urine collected during 2008–2016; 296 participants provided repeat urine and updated exposure information in 2025. Detection frequencies, co-detection, and within-person detection patterns were characterized. Generalized estimating equations were used for stacked, repeated-measures analyses of factors associated with detection of aminomethylphosphonic acid (AMPA), glyphosate, 2,4-dichlorophenoxyacetic acid (2,4-D), and any of these three. Prospective-only analyses evaluated more detailed agricultural and recent exposure measures.

**Results:** Glyphosate, AMPA, and 2,4-D were detected in 7.7%, 6.2%, and 4.3% of retrospective specimens and 5.4%, 3.1%, and 4.1% of prospective specimens, respectively. Co-detection and persistent detection across the 9–17-year interval was rare. In repeated-measures models, greater fruit and vegetable intake, older age, and male sex were associated with higher 2,4-D detection. Lower household income was associated with lower AMPA detection, while afternoon/evening collection was associated with higher AMPA detection. In prospective analyses, working on field-crop agricultural land showed the strongest agricultural associations, particularly for 2,4-D and detection of any of the three pesticides. Associations were not seen with self-reported conventional versus organic produce consumption.

**Conclusions:** Urinary pesticide detections were generally infrequent in this Wisconsin population. Diet and direct agricultural activities may be more informative exposure pathways than residing near cropland or private well drinking-water characteristics.

## INTRODUCTION

Population-based pesticide biomonitoring has expanded considerably in Europe in recent years. The Human Biomonitoring for Europe initiative harmonized pesticide measurements across multiple adult populations and documented measurable exposure to glyphosate, aminomethylphosphonic acid (AMPA), pyrethroids, chlorpyrifos-related biomarkers, and other contemporary-use pesticides across several countries (H. Andersen et al., 2022; H. R. Andersen et al., 2022; Buekers et al., 2022; Gilles et al., 2021). In France, the nationally representative Esteban study measured biomarkers spanning herbicides, pyrethroids, organophosphates, carbamates, and organochlorines and provided population distributions and exposure determinants for numerous pesticides, including glyphosate and AMPA (Tagne-Fotso et al., 2023). More recently, Ottenbros and colleagues applied a broad urinary suspect-screening approach among adults in the Netherlands and Switzerland, detecting biomarkers representing numerous pesticides and demonstrating both co-occurrence and substantial variation in exposure within and between populations (I. B. Ottenbros et al., 2023). Subsequent population biomonitoring studies in Spain and rural Latvia have similarly demonstrated heterogeneous pesticide exposure profiles and identified diet, pesticide use, and other local characteristics as potential exposure determinants (Akūlova et al., 2026a, 2026b; López et al., 2025). Together, these studies demonstrate the value and feasibility of broad population-based pesticide biomonitoring but also underscore that exposure profiles are geographically specific and dependent on pesticide use, diet, residential characteristics, and other local exposure sources.

In the United States, nationally representative pesticide biomonitoring is conducted primarily through the Centers for Disease Control and Prevention (CDC) National Health and Nutrition Examination Survey (NHANES). These data provide critical reference information but have generally focused on selected pesticides or pesticide classes during specific survey cycles rather than a consistent, broad panel of contemporary-use compounds. For example, urinary glyphosate was measured in NHANES 2013–2014, with detectable concentrations in approximately 81% of the U.S. population aged six years and older (Ospina et al., 2022).

NHANES data have also demonstrated temporal increases in urinary 2,4-dichlorophenoxyacetic acid (2,4-D), corresponding with increasing agricultural use of 2,4-D in the United States (Freisthler et al., 2022). More recent analyses of NHANES 2015–2016 have used urinary biomarkers spanning multiple pesticide classes to evaluate dietary pesticide exposure associated with produce consumption (Temkin et al., 2025). These studies provide important national benchmarks, but they do not comprehensively characterize the broader set of pesticides currently used in U.S. agricultural settings, nor are they designed to characterize region-specific exposure patterns in populations living in areas of intensive agriculture.

This gap is particularly relevant to the U.S. Midwest, where corn, soybean, small-grain, and livestock agriculture may result in pesticide-use patterns and environmental exposure opportunities that differ from those represented by national averages. Earlier biomonitoring studies in Iowa and Minnesota demonstrated measurable urinary exposure to glyphosate, 2,4-D, atrazine, chloroacetanilide herbicides, chlorpyrifos, and other agriculturally relevant pesticides among farmers, pesticide applicators, and farm families (Acquavella et al., 2004; Alexander et al., 2007; Curwin et al., 2007). More recent studies have continued to demonstrate differences in pesticide exposure associated with farming and occupational pesticide use, including the U.S. Biomarkers of Exposure and Effect in Agriculture study and biomonitoring studies conducted in other intensive agricultural regions (Chang et al., 2024; DeSantiago et al., 2026). However, much of the U.S. agricultural biomonitoring literature remains based on occupationally exposed groups, farm families, pregnancy cohorts, or relatively small regional samples rather than population-based samples of adults that include both agricultural and non-agricultural exposure pathways. Recent European population studies have begun to fill this gap internationally, but comparable broad population-based pesticide biomonitoring remains limited in the United States.

An additional limitation is the scarcity of long-term repeated pesticide biomonitoring within the same individuals. Most national surveillance programs, including NHANES, rely on repeated cross-sectional samples and therefore can characterize population trends but not individual changes in exposure. Recent serial cross-sectional biomonitoring studies have similarly provided important information on temporal changes in population pesticide exposure but do not follow the same individuals over time (Jia et al., 2025). This distinction is important for nonpersistent pesticides, for which urinary concentrations may vary substantially over relatively short periods. In the Ottenbros study, within-person correlations between repeated seasonal samples were low, illustrating considerable temporal variability in urinary pesticide biomarkers (I. Ottenbros et al., 2023; I. B. Ottenbros et al., 2023). Long-term studies that combine population-based sampling, archived biospecimens, and repeat biomonitoring of the same participants remain uncommon, limiting understanding of how pesticide exposure patterns change within individuals over years and whether co-detection patterns and exposure determinants persist over time.

To address these gaps, we leveraged the Survey of the Health of Wisconsin (SHOW), a population-based cohort with substantial representation of rural Wisconsin adults and archived urine specimens collected during 2008– 2016, with repeat urine collection and updated exposure assessment in 2025. The objective of this study was to characterize urinary pesticide detections, evaluate temporal changes in exposure, including within-person changes over time, and identify factors associated with detection of the most observed pesticides.

## METHODS

### Study population

The study sample was recruited from the SHOW, a statewide population-based health examination survey modeled after the NHANES. SHOW sampling and methods have been described previously (Malecki et al., 2022). Briefly, address-based area probability sampling without replacement was used to recruit statewide samples of Wisconsin residents during 2008–2016 (n=5,353). Wave I (2008–2013) included annual statewide samples of adults aged 21–74 years (n=3,380), and Wave II (2014–2016) included a triannual statewide sample of adults aged ≥18 years (n=1,973). Participants completed interviewer- and self-administered questionnaires, objective health measurements, and biospecimen collection and consented to storage of survey data and biospecimens for future research and to be contacted for future ancillary studies.

### Eligibility and recruitment

SHOW adults aged ≥18 years were eligible for the present study if they had sufficient archived urine from their 2008–2016 SHOW examination, currently resided in a rural area or smaller urban cluster adjacent to a rural area, and resided within one of three agricultural study regions in Wisconsin. Residents of larger urbanized areas were excluded, leaving a total of 1,247 eligible adults. Eligible adults were contacted by mail and, when available, email and invited to complete a brief screening questionnaire using University of Wisconsin-Madison’s (UW) REDCap (Research Electronic Data Capture) data collection tool online or by telephone with study staff (Harris et al., 2019). The screener confirmed current residence and that the location met the eligibility criteria above. Upon completion, eligible participants were provided with available community site locations and appointment times to schedule their appointment within the REDCap tool.

### Prospective survey interview and specimen collection

Prospective study visits were conducted in or near communities where eligible participants resided, including at local health departments, libraries, churches, senior centers, and other community sites. Sites held appointments for one day up to 14 days depending on the numbers of eligible participants in the vicinity and were conducted over a 5 month period, from June to November of 2025. Visits lasted approximately 40 minutes and included informed consent, an interviewer-administered questionnaire entered directly into REDCap, and collection of a clean-catch spot urine specimen. Participants were compensated $40 for completing the study. The Health Sciences Institutional Review Board of the University of Wisconsin-Madison gave ethical approval for this work.

### Baseline survey measures

Baseline participant characteristics were obtained from the SHOW examination corresponding to each participant’s 2008–2016 urine collection. Measures included demographic and socioeconomic characteristics, residential and drinking-water characteristics, diet, household pesticide use, and geocoded residential characteristics.

#### Demographic and socioeconomic characteristics

Age was defined at the time of specimen collection. Sex, race, Hispanic/Latino ethnicity, educational attainment, and household income were self-reported. Race was categorized as White versus non-White for analysis due to too few non-White participants, and Hispanic/Latino ethnicity was not retained in analysis due to too few Hispanic/Latino adults. Education was harmonized as less than high school, high-school graduate/General Educational Development (GED) or some college, and bachelor’s degree or higher. For prospective-only analyses, education was additionally categorized as less than a bachelor’s degree versus bachelor’s degree or higher to reduce sparse cells.

Annual household income was collected in ordered categories. The midpoint of the reported income category was used for analyses requiring an estimated household income and was categorized as <$75,000 versus ≥$75,000. Household income and the number of persons supported by that income were also used to calculate the poverty-income ratio based on the U.S. Department of Health and Human Services annual Federal Poverty Level (FPL) guidelines (U.S. Department of Health and Human Services, 2024). The poverty-income ratio was calculated as household income divided by the poverty guideline for the corresponding household size; values were multiplied by 100 when expressed as percent FPL. Participants were categorized as ≤200% versus >200% FPL.

#### Residential and drinking-water characteristics

Participants reported whether the primary source of water used for drinking at home was a private well or community/municipal water supply, whether they filtered their water, and what type of filtration system they used. Water filtration was harmonized and analyzed as "uses water filter" or "does not use water filter", with no filter use as the reference group. “Uses water filter” was defined as use of an under-the-sink, faucet, or pitcher carbon filter or a reverse osmosis system. Water softeners and aerators were not considered water filters as they do not remove harmful contaminants measured in this study.

Residential urbanicity was assigned from the geocoded home address using the 2010 U.S. Census urban-rural classification. Under the 2010 Census definition, urbanized areas contained ≥50,000 residents, urban clusters contained 2,500–49,999 residents, and all population and territory outside Census-defined urban areas was classified as rural (U.S. Census Bureau, 2020). Larger urbanized areas were excluded from the study eligibility frame; thus, the analytic sample included rural residents and residents of smaller urban clusters.

Neighborhood socioeconomic context was characterized using the Area Deprivation Index (ADI) from the Neighborhood Atlas (Kind and Buckingham, 2018). The ADI is a census-block-group measure incorporating 17 indicators of income, education, employment, and housing conditions; higher values indicate greater neighborhood socioeconomic deprivation. Wisconsin state ADI deciles were linked to participant addresses, using the 2015 ADI for retrospective observations. State deciles range from 1 to 10, with 10 representing the greatest relative deprivation within Wisconsin. ADI was evaluated continuously per one-decile increase and categorically. The primary binary comparison grouped deciles 1–6 versus 7–10, thereby contrasting residents of the 40% most deprived Wisconsin block groups with those in deciles 1–6. Alternative analyses classified ADI as deciles 1–3, 4–7, and 8–10.

Percentage of cropland within a one-mile buffer of the participant’s geocoded residential address was calculated using ESRI ArcGIS using WISCLAND land cover data. The land cover data came from the Wisconsin Department of Natural Resources, which derived land cover data from satellite imagery acquired from the Landsat 5 Thematic Mapper, Landsat 7 Enhanced Thematic Mapper, and Landsat 8 Operational Land Imager between 2010 and 2014 (WDNR & UW-Madison, 2016). Cropland was evaluated continuously per 20-percentage-point increase and categorically as <40% versus ≥40% of land within one mile of the residence. Participants were additionally assigned to the southern, western, or central Wisconsin study region based on county of residence.

#### Diet

Dietary assessment differed across the two SHOW baseline waves. During 2008–2013, SHOW used a questionnaire modified from the NutritionQuest Block Fat/Sugar/Fruit/Vegetable Screener, which generated separate estimates of usual fruit and vegetable intake (Lalonde et al., 2008). Beginning in 2014, SHOW transitioned to a National Cancer Institute (NCI) dietary screener that generates a combined fruit-and-vegetable daily intake estimate (Millen et al., 2006). SHOW documentation describes the use of validated dietary screeners as part of its self-administered questionnaire battery. Because fruit and vegetable intake was calculated separately under the Block-based instrument but jointly under the NCI instrument, fruit and vegetable intake from 2008–2013 was combined to create a common measure that could be harmonized with the 2014–2016 estimate. Harmonized fruit and vegetable intake was evaluated continuously, per 2-serving/day increase. Participants were also asked about consumption of organic food. For the retrospective survey, this was assessed as whether participants ate organic food (yes/no).

#### Household pesticide use

Participants reported whether pesticides, including insecticides, fungicides, or herbicides, had been used inside and outside the home during the previous 12 months. Indoor and outdoor pesticide use were each evaluated as binary yes/no measures, and an additional indicator identified participants reporting either indoor or outdoor use.

The format used to assess frequency of outdoor pesticide application changed between SHOW waves. From 2008–2013, participants reporting outdoor pesticide use entered the number of times pesticides had been applied during the previous 12 months. Beginning in 2014, frequency was collected using categorical response options: 0 times, 1 time, 2–3 times, or ≥4 times. To harmonize the measure across study years, integer responses from 2008–2013 were recoded to these same four categories. Indoor pesticide application frequency was assessed and harmonized in the same manner. Outdoor and indoor application frequency was collected separately. Participants additionally reported who applied the pesticides and, when applicable, the method of application for both indoor and outdoor pesticides. All SHOW baseline survey instruments can be found here: https://data.show.wisc.edu/data/codebooks/

### Prospective follow-up survey measures

The 2025 prospective follow-up survey repeated core SHOW demographic, socioeconomic, housing, drinking-water, diet, and household pesticide-use measures using the same or harmonizable question wording and response categories when possible to support longitudinal analyses. Primary drinking-water source and water filtration were asked how they were in the baseline survey and harmonized for analysis. Drinking water source was again classified as private well versus community/municipal water. The prospective survey also retained the baseline 12-month indoor and outdoor pesticide-use questions, which were harmonized for analysis.

The prospective questionnaire also included additional measures designed to characterize agricultural activities and pesticide exposures occurring closer to urine collection. Additional prospective questions assessed whether participants currently lived on or adjacent to agricultural land, currently worked on agricultural land, and worked directly in planting or harvesting. The items also asked what type of agricultural land participants lived on or near or worked directly on or with (e.g. corn, soybean, legumes, small grain crops, vegetables, fruits, pasture or forage, small livestock). Recent pesticide contact was assessed by asking whether participants had handled or used pesticides during paid or unpaid work in the previous three months and whether they had contacted vegetation or soil treated with pesticides during the previous three months, including during lawn maintenance or gardening. These measures were available only for prospective analyses.

Diet was modified in the prospective survey to better capture intake during the short exposure window relevant to urinary pesticide biomarkers. Participants separately reported the number of times they had consumed fresh fruits and fresh vegetables during the previous three days. Fruit intake, vegetable intake, and their sum were therefore evaluated separately in prospective-only analyses; for longitudinal analyses, fruit and vegetable intake was combined to correspond with the harmonized baseline measure.

The prospective survey also asked how often participants consumed foods described as organic, pesticide-free, or chemical-free using the responses: always, most of the time, sometimes, rarely, or never. For analysis, this was harmonized with the baseline yes/no organic-food item by classifying always/most of the time as yes and sometimes/rarely/never as no. The prospective survey is available in Supplementary Materials.

Residential contextual measures were updated for the participant’s follow-up address. The 2023 Wisconsin state ADI was used for prospective observations, allowing state-decile measures to be harmonized with the 2015 ADI used retrospectively. Cropland within one mile, Census urban-rural classification, and study region were assigned using the same definitions as in the retrospective period.

## Specimen collection and handling

At the prospective visit, participants provided a clean-catch spot urine specimen. Date and time of collection were recorded. Specimens were aliquoted onsite and stored in locked portable −80°C freezers until transport to Madison on dry ice, generally weekly or biweekly, where they were transferred to long-term −80°C storage. Retrospective analyses used archived spot urine collected during participants’ original 2008–2016 SHOW examinations. For analysis, collection season was categorized as April–October versus November–March. Time of collection was initially categorized as 6:00 AM–11:59 AM, 12:00 PM–5:59 PM, or 6:00 PM–9:00 PM; afternoon and evening collections were combined for primary analyses and compared with morning collections.

### Laboratory methods for urinary pesticide detection

Urinary analyses of 28 pesticide-related analytes were conducted by the Wisconsin State Laboratory of Hygiene (WSLH). The analyte panel was selected based on several considerations, including: (1) relevance to pesticide use in Wisconsin and Midwestern agricultural systems; (2) pesticides previously identified through Wisconsin groundwater and surface-water monitoring; (3) representation of multiple pesticide classes and both commonly used and emerging pesticides of concern; (4) inclusion of selected parent compounds and their metabolites or degradation products, including glyphosate and aminomethylphosphonic acid (AMPA); and (5) the feasibility of developing reliable urinary measurement methods given available analytical standards and reference materials. Wisconsin Department of Agriculture, Trade and Consumer Protection (DATCP) surveillance provided an important basis for identifying pesticides of state relevance. Recent statewide and targeted groundwater monitoring has identified multiple herbicides, insecticides, fungicides, and pesticide metabolites in Wisconsin drinking-water wells, while surface-water monitoring has similarly documented frequent detection of agricultural pesticides and their degradation products (DATCP, 2024). Neonicotinoid insecticides were specifically included because of their widespread agricultural use and their identification as emerging contaminants of concern in Wisconsin groundwater.

The final panel consisted of <u>20 herbicide or herbicide-related analytes, seven insecticides, and one fungicide</u>. Herbicides included several major classes used in Wisconsin agriculture, including phenoxy herbicides, chloroacetanilides, triazines, glyphosate and AMPA, and other current-use herbicides. The insecticide panel included four neonicotinoids (acetamiprid, clothianidin, imidacloprid, and thiamethoxam), two diamide insecticides (chlorantraniliprole and cyantraniliprole), and the carbamate insecticide carbaryl; metalaxyl was included as a fungicide. The complete panel, pesticide classification, analyte-specific limits of detection, and detection frequencies are provided in Supplementary Tables 1-4.

Urine specimens were analyzed at the Wisconsin State Laboratory of Hygiene (WSLH) for pesticide biomarkers using ultra-high-performance liquid chromatography coupled with electrospray ionization tandem mass spectrometry (UPLC-ESI-MS/MS; Agilent Technologies 1290 Infinity II LC coupled to a SCIEX 4500 tandem mass spectrometer). Two analytical methods were used: a 26-analyte pesticide biomarker panel comprising of 22 analytes measured in positive-ionization mode and four analytes measured in negative-ionization mode, and a separate two-analyte method for glyphosate and aminomethylphosphonic acid (AMPA). For the 26-analyte panel, urine specimens were extracted with methanol and centrifuged; the resulting supernatant was collected, and evaporated to approximately 200 µL prior to analysis. Chromatographic separation was performed using an Acquity UPLC BEH C18 column, followed by multiple-reaction monitoring (MRM) using analyte-specific precursor-to-product ion transitions. Isotopically labeled internal standards were incorporated where available. Analyte-specific MRM transitions, retention times, and other instrument parameters are provided in Supplementary Table 2. Glyphosate and AMPA were analyzed separately following derivatization with 9-fluorenylmethyl chloroformate (FMOC-Cl) and solid-phase extraction (SPE) cleanup using a Phenomenex Strata Si-1 silica SPE plate (55 µm, 70 Å, 100 mg). Extracts were evaporated to dryness, reconstituted in water, and chromatographically separated using a Synergi Fusion-RP column prior to tandem mass spectrometric detection.

Both pesticide biomarker exposure methods were developed and validated by WSLH as quantitative high-complexity laboratory-developed tests in accordance with applicable CLIA regulatory standards and College of American Pathologists (CAP) accreditation requirements for clinical laboratory testing. Method validation included evaluation of analytical measurement range, accuracy, within-run and between-run precision, analytical specificity, and carryover. Routine quality assurance and quality control procedures included multipoint calibration using verified reference materials, isotopically labeled internal standards, matrix-matched reference materials and quality-control samples, and controls spanning the analytical measurement range. Method performance was additionally evaluated through alternative proficiency testing procedures. Analyte-specific limits of detection (LODs) were established during method validation and are reported in Supplementary Tables 2 and 3. A pesticide biomarker was considered detected when the measured urinary concentration exceeded the analyte-specific LOD.

Urinary creatinine was quantified using the QuantiChrom Creatinine Assay Kit (BioAssay Systems) with absorbance measured using an Agilent BioTek Epoch 2 microplate spectrophotometer. Creatinine measurements were used to calculate creatinine-corrected pesticide biomarker concentrations for descriptive concentration summaries. Pesticide biomarker detection status was determined using the uncorrected urinary concentration relative to the corresponding laboratory LOD.

### Data and statistical analysis

Study period was defined as retrospective (2008–2016) versus prospective (2025). For analysis combining retrospective and prospective observations, only measures that could be meaningfully harmonized across study periods were included. Measures unique to the 2025 questionnaire—including residence on or adjacent to agricultural land, agricultural work, direct planting or harvesting, occupational pesticide handling, contact with pesticide-treated vegetation or soil, and separate three-day fruit and vegetable intake—were evaluated only in prospective analyses.

Participant characteristics were summarized separately for the retrospective and prospective study periods using means and standard deviations for continuous measures and frequencies and percentages for categorical measures. Characteristics of adults who completed prospective follow-up were compared with the full eligible recruitment sample using chi-square tests. Detection frequencies were calculated for each of the 28 urinary pesticide analytes separately for retrospective and prospective specimens. For descriptive summaries of pesticide concentrations, urinary concentrations were corrected for creatinine and summarized using geometric means, medians, and ranges. Detection status, however, was based solely on whether the measured urinary concentration exceeded the laboratory limit of detection and was not creatinine-adjusted. For participants with specimens available at both time points, detection patterns were additionally classified as detected retrospectively only, prospectively only, at both time points, or at neither time point.

Because most analytes were rarely or never detected, regression analyses were limited to the three pesticides detected most frequently and in both study periods: AMPA, glyphosate, and 2,4-D. For each analyte, the primary outcome was detection above the LOD. A fourth binary outcome indicated detection of any of these three analytes. Analyses therefore evaluated predictors of pesticide detection rather than urinary pesticide concentration.

The primary analyses combined retrospective and prospective observations in a stacked longitudinal dataset. Generalized estimating equations (GEE) with a logit link and binomial distribution were used to estimate odds ratios (ORs) and 95% confidence intervals (CIs) while accounting for within-participant correlation among adults contributing observations at both time points. An exchangeable working correlation structure was specified, assuming a common correlation between repeated observations from the same participant, with robust (sandwich) standard errors. All stacked, repeated measures models included study period (retrospective versus prospective) to account for differences in detection between collection periods. Separate logistic regression models were also estimated for retrospective and prospective observations to examine whether associations differed by study period, examine robustness of associations seen, and evaluate exposure measures available only from the prospective questionnaire.

Potential predictors included demographic and socioeconomic characteristics, residential and geographic characteristics, drinking-water source and filtration, diet, household pesticide use, and specimen collection characteristics. Prospective analyses additionally evaluated agriculture near residence and recent occupational or nonoccupational pesticide contact. General agricultural residence and work were evaluated as separate binary exposures. Crop-specific indicators were then derived and analyzed separately for field crops (corn, soybeans, or small grains), specialty crops (vegetables or fruits), and pasture/forage, with separate variables for living on or adjacent to and working on each land type. These crop-specific analyses were considered exploratory because they were based on smaller exposed subgroups and, consequently, often produced less precise estimates. Univariate associations with each pesticide outcome were first evaluated separately. For the stacked dataset, univariate models included the predictor of interest and study period; prospective- and retrospective-only univariate models included the predictor of interest alone. ORs and 95% CIs from these models were used to characterize the direction, magnitude, and precision of associations and to inform development of parsimonious multivariable models.

Multivariable models included age and sex a priori, and stacked models additionally included study period. Given the limited number of pesticide detections, the number of additional predictors included in each model was restricted to avoid overparameterization. Candidate predictors were evaluated based on the magnitude and precision of univariate associations, consistency of associations across study periods, relevance to plausible pesticide exposure pathways, and correlation with other candidate predictors. Final models were selected to retain the most informative and parsimonious set of predictors for each outcome. Prospective multivariable models were limited to age, sex, and no more than two or three additional predictor constructs because of the smaller sample size and number of detections.

Analyses were conducted using complete observations for variables included in each model. Statistical analyses were performed in R version 4.6.0 (R Foundation for Statistical Computing, Vienna, Austria). Statistical tests were two-sided. Because analyses of potential exposure determinants were intended to identify and characterize patterns of association rather than test a single prespecified hypothesis, interpretation emphasized effect estimates, 95% CIs, consistency across analyses, and biological plausibility rather than statistical significance alone.

## RESULTS

### Study population

Among 1,248 SHOW participants who were invited to prospective follow-up, 391 responded to recruitment (31.3%), 329 scheduled a study appointment (26.4%), and 296 completed the prospective study visit (23.7%). An additional 304 eligible adults with stored baseline urine were included for the retrospective analytic sample, totaling 600 adults with urine specimens collected during 2008–2016, of whom 296 completed prospective follow-up and provided a second urine specimen in 2025 (Table 1; Figure 1). At baseline, participants had a mean age of 51.9 years (Standard Deviation (SD), 14.7); at follow-up, mean age was 66.8 years (SD, 12.5), reflecting the longitudinal study design. Approximately 58% were female at both timepoints, and the sample was predominantly White. Most participants lived in rural areas (86.8% at baseline and 81.1% at follow-up), and private wells were the primary drinking-water source for 79.5% and 72.6%, respectively. The follow-up sample was more highly educated and had higher household income than the baseline sample; 41.6% had a bachelor’s degree or higher and 45.9% had household income ≥$75,000 in 2025. Retrospective urine collections occurred throughout the year, although 61.0% were collected during April–October; 60.2% were collected between 6:00 AM and 11:59 AM, 28.7% between 12:00 PM and 5:59 PM, and 11.2% between 6:00 PM and 9:00 PM. In contrast, prospective collections were intentionally concentrated during the 2025 growing and harvest season, with 99.0% collected during April–October; approximately half were collected in the morning (50.0%) and half between 12:00 PM and 5:59 PM (49.0%).

**Figure 1.**
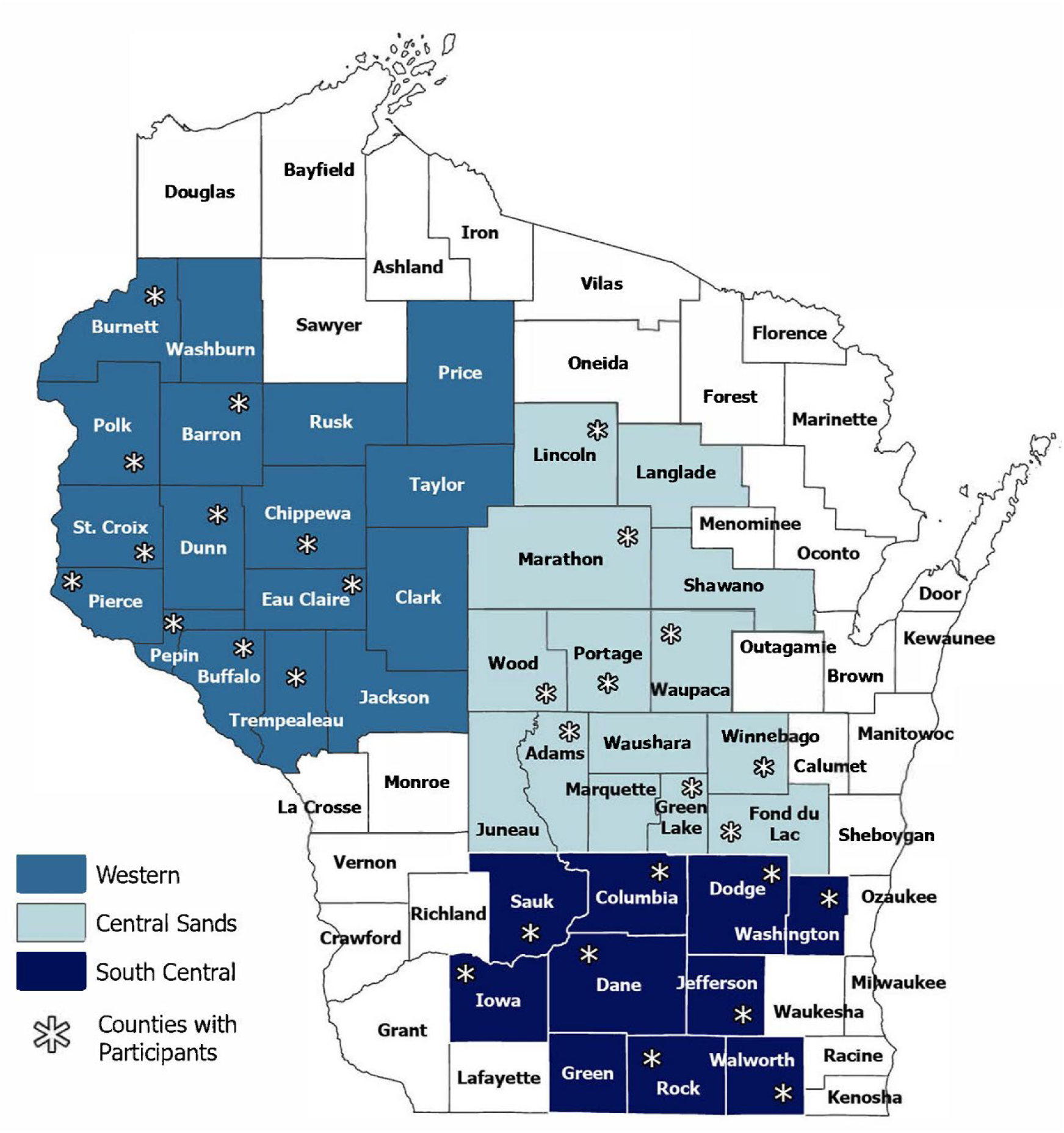
Wisconsin Rural Health Study recruitment regions and counties represented in the prospective study sample. Map shows the three Wisconsin study regions (Western, Central Sands, and South Central) used for prospective recruitment in 2025. Asterisks indicate counties with at least one participant who completed the prospective study visit.

**Table 1.** Characteristics of the baseline study sample (n=600) who enrolled in 2008-2016, and a subset who followed-up in 2025 (n=296).

| Characteristic |  | Baseline sample (n=600) |  | Follow-up sample (n=296) |  |
| --- | --- | --- | --- | --- | --- |
|  |  | n | Col % | n | Col. % |
| **DEMOGRAPHICS** |  |  |  |  |  |
| Age | Mean (SD) | 51.9 | 14.7 | 66.8 | 12.5 |
| Age |  |  |  |  |  |
|  | 18-35 | 103 | 17.2 | 2 | 0.7 |
|  | 35-49 | 133 | 22.2 | 33 | 11.2 |
|  | 50-64 | 232 | 38.7 | 66 | 22.3 |
|  | 65 or older | 132 | 22.0 | 195 | 65.9 |
| Sex |  |  |  |  |  |
|  | Male | 255 | 42.5 | 124 | 41.9 |
|  | Female | 345 | 57.5 | 172 | 58.1 |
| Race |  |  |  |  |  |
|  | White | 574 | 95.7 | 289 | 97.6 |
|  | Non-White | 25 | 4.2 | 7 | 2.4 |
|  | Missing | 1 | 0.17 |  |  |
| Federal Poverty Level (FPL) |  |  |  |  |  |
|  | Below 200% FPL | 123 | 20.5 | 55 | 18.6 |
|  | At or above 200% FPL | 460 | 76.7 | 235 | 79.4 |
|  | Missing | 17 | 2.8 | 6 | 2.0 |
| Household Income |  |  |  |  |  |
| | Below \$75,000 | 368 | 61.3 | 153 | 51.7 |
| | At or above \$75,000 | 214 | 35.7 | 136 | 45.9 |
|  | Missing | 18 | 3 | 7 | 2.4 |
| Highest Educational Attainment |  |  |  |  |  |
|  | Less than HS/GED | 143 | 23.8 | 5 | 1.7 |
|  | HS/GED or Some college | 247 | 41.2 | 168 | 56.8 |
|  | Bachelor's or more | 210 | 35.0 | 123 | 41.6 |
| **GEOGRAPHY** |  |  |  |  |  |
| Wisconsin Region |  |  |  |  |  |
|  | South | 153 | 23.8 | 62 | 21.0 |
|  | West | 209 | 34.8 | 106 | 33.8 |
|  | Central | 238 | 39.7 | 128 | 43.2 |
| Census Urban / Rural |  |  |  |  |  |
|  | Urban Cluster | 79 | 13.2 | 56 | 18.9 |
|  | Rural | 521 | 86.8 | 241 | 81.1 |
| Agriculture <1 mile of home |  |  |  |  |  |
|  | ≥40% | 214 | 35.7 | 211 | 71.3 |
|  | <40% | 386 | 64.3 | 85 | 28.7 |
| Area Deprivation Index (ADI) |  |  |  |  |  |
|  | Low deprivation 1-3 | 191 | 31.8 | 92 | 31.1 |
|  | Moderate 4-7 | 334 | 55.7 | 158 | 53.4 |
|  | High deprivation 8-10 | 74 | 12.3 | 40 | 13.5 |
|  | Missing (low population /housing) | 1 | 0 | 6 | 2 |
| **DIET & HOME** |  |  |  |  |  |
| Drinking water source |  |  |  |  |  |
|  | Private well | 477 | 79.5 | 215 | 72.6 |
|  | Municipal | 103 | 17.2 | 79 | 26.7 |
|  | Don't know or missing | 20 | 2.5 | 2 | 0.7 |
| Water filtration (pitcher, carbon, RO) |  |  |  |  |  |
|  | Yes | 163 | 27.2 | 108 | 36.5 |
|  | No | 419 | 69.8 | 141 | 47.6 |
|  | Don't know or missing | 18 | 3.0 | 47 | 15.9 |
| Daily Fruit and Veggie Intake |  |  |  |  |  |
|  | < 3.5 cups | 452 | 75.3 | 191 | 64.5 |
|  | ≥ 3.5 cups | 127 | 21.2 | 99 | 33.5 |
|  | Missing | 21 | 3.5 | 6 | 2.0 |
| **Eat organic** |  |  |  |  |  |
|  | Yes | 169 | 28.2 | 94 | 31.8 |
|  | No | 235 | 39.2 | 199 | 67.2 |
|  | *Missing* | *1* | *0.2* | *3* | *1.0* |
|  | *Not asked* | *195* | *32.5* |  |  |
| **Work with pesticides** |  |  |  |  |  |
|  | Yes |  |  | 32 | 20.7 |
|  | No |  |  | 123 | 79.4 |
| **Use pesticides INSIDE the home** |  |  |  |  |  |
|  | Yes | 176 | 29.3 | 165 | 55.7 |
|  | No | 397 | 66.2 | 131 | 44.3 |
|  | *Missing or Don't know* | *27* | *4.5* |  |  |
| **Use pesticides OUTSIDE the home** |  |  |  |  |  |
|  | Yes | 418 | 69.7 | 212 | 71.6 |
|  | No | 143 | 23.8 | 82 | 27.7 |
| **OUTSIDE pesticide application (times per year)** |  |  |  |  |  |
|  | 0 times | 143 | 23.8 | 82 | 27.7 |
|  | 1 time | 122 | 20.3 | 60 | 20.3 |
|  | 2-3 times | 185 | 30.8 | 92 | 31.1 |
|  | 4 or more times | 104 | 17.3 | 55 | 18.6 |
|  | *Missing or Don't know* | *46* | *7.7* | *7* | *2.4* |
| **OTHER** |  |  |  |  |  |
| **Smoking status** |  |  |  |  |  |
|  | Current | 72 | 12.0 | 13 | 4.4 |
|  | Former or never | 512 | 85.3 | 282 | 95.6 |
|  | *Missing* | *16* | *2.7* |  |  |
| **Year data/specimen collected** |  |  |  |  |  |
|  | 2025 | 0 | 0.0 | 296 | 100.0 |
|  | 2014-2016 | 237 | 40.7 | 0 | 0.0 |
|  | 2011-2013 | 205 | 35.2 | 0 | 0.0 |
|  | 2008-2010 | 141 | 24.2 | 0 | 0.0 |
| **Urinary Creatinine** |  |  |  |  |  |
|  | Dilute/low (<30 mg/dL) | 72 | 12.0 | 66 | 22.3 |
|  | Normal to high (≥ 30 mg/dL) | 528 | 88.0 | 230 | 77.7 |
| **Season specimen collected** |  |  |  |  |  |
|  | Winter (Nov-Mar) | 198 | 33.0 | 3 | 1.0 |
|  | Spring, Summer, Fall (Apr-Oct) | 366 | 61.0 | 293 | 99.0 |
|  | *Missing* | *36* | *6* |  |  |
| **Time of day specimen collected** |  |  |  |  |  |
|  | 6a-11:59a | 361 | 60.2 | 148 | 50.0 |
|  | 12p-5:59p | 172 | 28.7 | 145 | 49.0 |
|  | 6p-9p | 67 | 11.2 | 3 | 1.0 |

Compared with the full group of eligible SHOW participants, those completing prospective follow-up were substantially older and were more likely to have higher educational attainment and household income and to live in less socioeconomically disadvantaged areas (Supplementary Table S1). Prospective participants were also somewhat more likely to reside in areas with greater surrounding cropland, whereas sex distribution and rural residence were similar between participants and the overall eligible sample. These differences indicate some selection toward older and more socioeconomically advantaged participants in the prospective sample.

### Urinary pesticide detection and longitudinal patterns

Pesticide detections were uncommon across both study periods (Table 2). Glyphosate was the most frequently detected analyte, detected in 7.7% of retrospective and 5.4% of prospective specimens, followed by AMPA (6.2% and 3.1%, respectively) and 2,4-D (4.3% and 4.1%). Sulfentrazone was detected in 1.0% of retrospective specimens and 0.3% of prospective specimens; triclopyr and imidacloprid were each detected once retrospectively, and 2-methyl-4-chlorophenoxyacetic acid (MCPA) was detected once prospectively. None of the other pesticides tested were detected.

**Table 2.** Frequency of detection and descriptive statistics for 28 pesticides in urine among retrospective (n=600) and prospective (n=296) participant samples. Geomeans, median and range urinary pesticide concentations are creatinine-corrected and reported in µg/g.

| Analyte | Limit of Detection (LOD) µg/L | Retrospective (n=600) |  |  |  | Prospective (n=295) |  |  |  |
| --- | --- | --- | --- | --- | --- | --- | --- | --- | --- |
|  |  | n (%) above LOD | Geomean (µg/g) | Min (µg/g) | Max (µg/g) | n (%) above LOD | Geomean (µg/g) | Min (µg/g) | Max (µg/g) |
| Herbicides |  |  |  |  |  |  |  |  |  |
| Glyphosate | 0.5 | 45 (7.7) | 0.48 | 0.59 | 6.16 | 16 (5.4) | 0.68 | 0.88 | 4.25 |
| 2,4-Dichlorophenoxyacetic acid (2,4-D) | 1.0 | 25 (4.3) | 0.91 | 1.14 | 15.92 | 12 (4.1) | 1.33 | 1.72 | 18.88 |
| Aminomethylphosphonic acid (AMPA) | 0.5 | 36 (6.2) | 0.46 | 0.59 | 5.33 | 9 (3.1) | 0.65 | 0.85 | 5.85 |
| Sulfentrazone | 0.2 | 6 (1.0) | 0.17 | 0.23 | 2.13 | 1 (0.3) |  |  | 1.70 |
| 2-Methyl-4-chlorophenoxyacetic acid | 0.5 | 0 (0) |  |  |  | 1 (0.3) |  |  | 4.25 |
| Triclopyr | 25.0 | 1 (0.2) |  |  | 266.63 | 0 (0) |  |  |  |
| Insecticides |  |  |  |  |  |  |  |  |  |
| Imidacloprid | 2.5 | 1 (0.2) |  |  | 26.66 | 0 (0) |  |  |  |

Co-detection was also uncommon (Figure 2; Supplementary Table S4). Glyphosate and AMPA were the predominant co-detection pattern, occurring in 2.0% of retrospective specimens and 1.0% of prospective specimens. Other combinations occurred in ≤0.3% of specimens. Among the 296 participants with specimens at both timepoints, persistence of individual pesticide detections over the approximately 9–17-year interval was rare (Figure 3; Supplementary Table S5). Glyphosate was detected only retrospectively in 18 participants, at both timepoints in 2, and only prospectively in 14. Corresponding counts were 16, 1, and 8 for AMPA and 10, 3, and 9 for 2,4-D. Sulfentrazone, triclopyr, and MCPA were each detected in no more than one participant at either timepoint, with no persistent detections.

**Figure 2.**
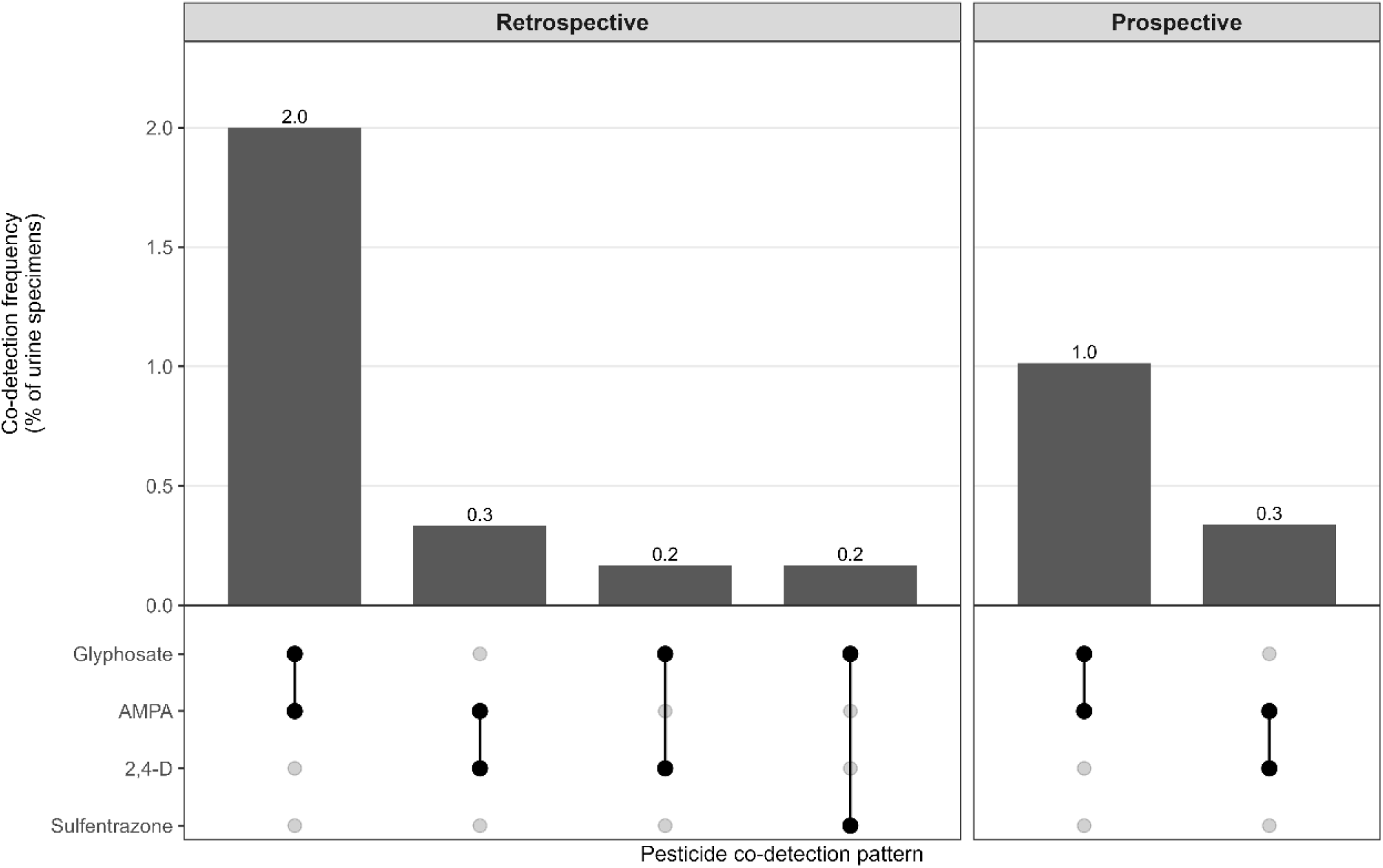
Co-detection patterns of urinary pesticides by study period. Bars show the percentage of urine specimens with each observed combination of two or more detected pesticides among retrospective specimens collected during 2008–2016 (n=600) and prospective specimens collected in 2025 (n=296). The matrix below each panel identifies the pesticides comprising each co-detection pattern. Specimens with no pesticide detected or only one pesticide detected are not shown. AMPA, aminomethylphosphonic acid; 2,4-D, 2,4-dichlorophenoxyacetic acid.

**Figure 3.**
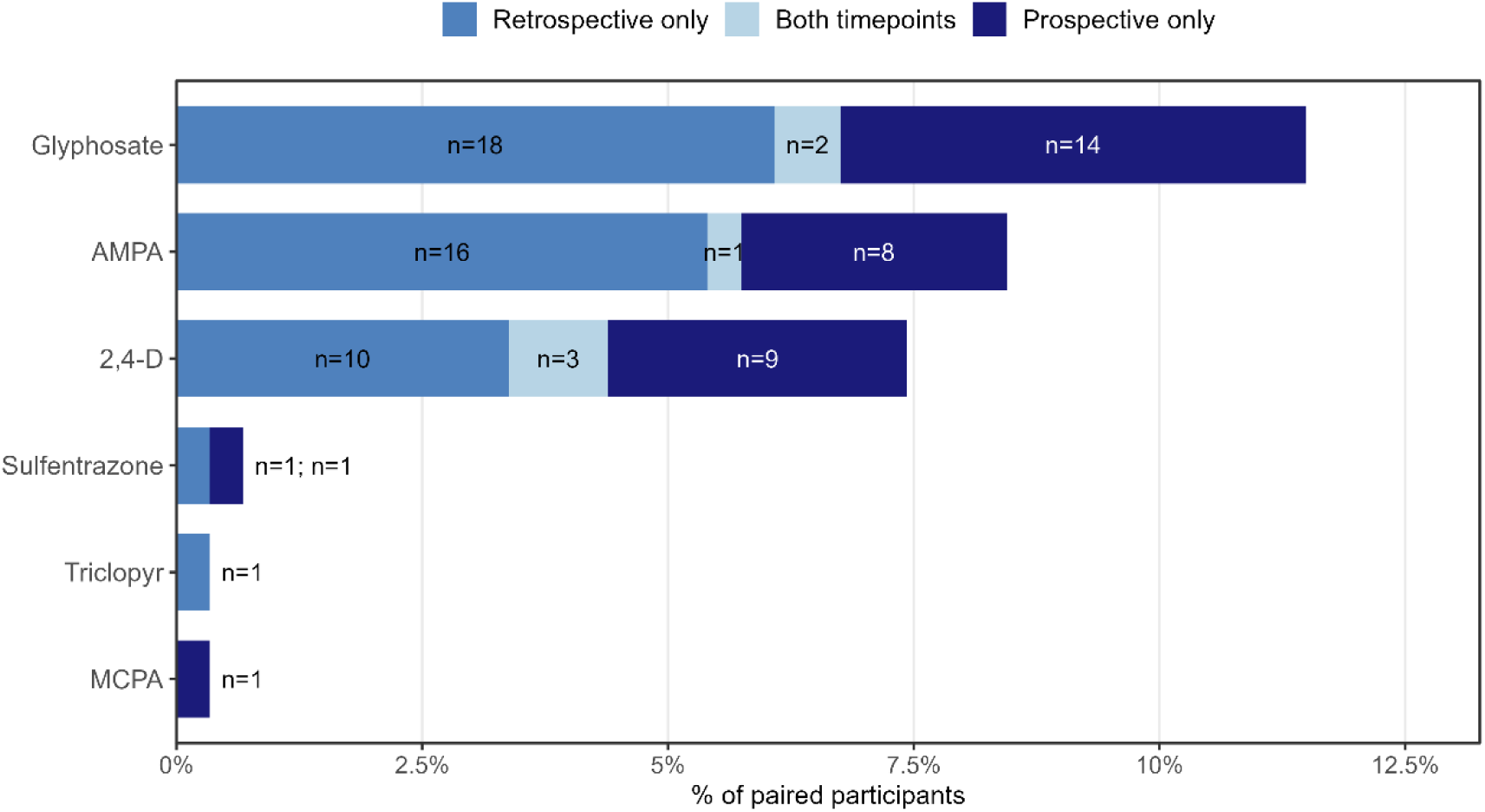
Longitudinal patterns of urinary pesticide detection among participants with paired specimens. Among 296 participants with both retrospective (2008–2016) and prospective (2025) urine specimens, bars show the percentage with each pesticide detected only retrospectively, at both timepoints, or only prospectively. Labels indicate the number of participants in each detection pattern. Only pesticides detected in at least one paired specimen are shown. AMPA, aminomethylphosphonic acid; 2,4-D, 2,4-dichlorophenoxyacetic acid; MCPA, 2-methyl-4-chlorophenoxyacetic acid.

### Factors associated with pesticide detection

Univariate associations from the stacked, repeated-measures analyses and the separate retrospective and prospective analyses were generally directionally consistent, although estimates were less precise in prospective-only analyses because of the smaller sample and lower numbers of detections (Supplementary Tables S6–S8; Supplementary Figure S1). Several expected exposure-related variables were not strongly associated with pesticide detection. Drinking-water filtration showed little consistent association with any outcome. Outdoor household pesticide use generally trended toward higher odds of glyphosate detection and detection of any of the three primary pesticides, with increasing frequency of outdoor application showing similar positive patterns, but confidence intervals were wide. Indoor pesticide use was not consistently associated with higher detection and, prospectively, was inversely associated with glyphosate detection. Surrounding cropland within one mile of the residence was also not consistently associated with detection in the stacked or phase-specific analyses.

Collection characteristics showed a more distinct pattern. Afternoon/evening compared with morning collection was associated with higher odds of AMPA detection in the stacked analysis, with a similar association in the retrospective data; evening collection was also associated with higher glyphosate detection retrospectively. In contrast, collection season was not strongly associated with detection in the retrospective or stacked analyses. Because nearly all prospective specimens were collected during April–October, prospective data provided little ability to evaluate seasonal differences independently.

In final multivariable repeated-measures models (Figure 4), several associations remained after adjustment. Household income <$75,000 was associated with lower odds of AMPA detection (OR, 0.46; 95% CI, 0.24– 0.86), while specimens collected from 12 PM–9 PM had approximately twice the odds of AMPA detection compared with morning specimens (OR, 2.10; 95% CI, 1.12–3.94). For glyphosate, compared with participants with less than a high-school education, odds of detection were lower among those with a high-school degree/GED or some college (OR, 0.40; 95% CI, 0.20–0.83) and those with a bachelor’s degree or higher (OR, 0.38; 95% CI, 0.17–0.85). Private-well use was also inversely associated with glyphosate detection, although the confidence interval included the null (OR, 0.57; 95% CI, 0.30–1.07).

**Figure 4.**
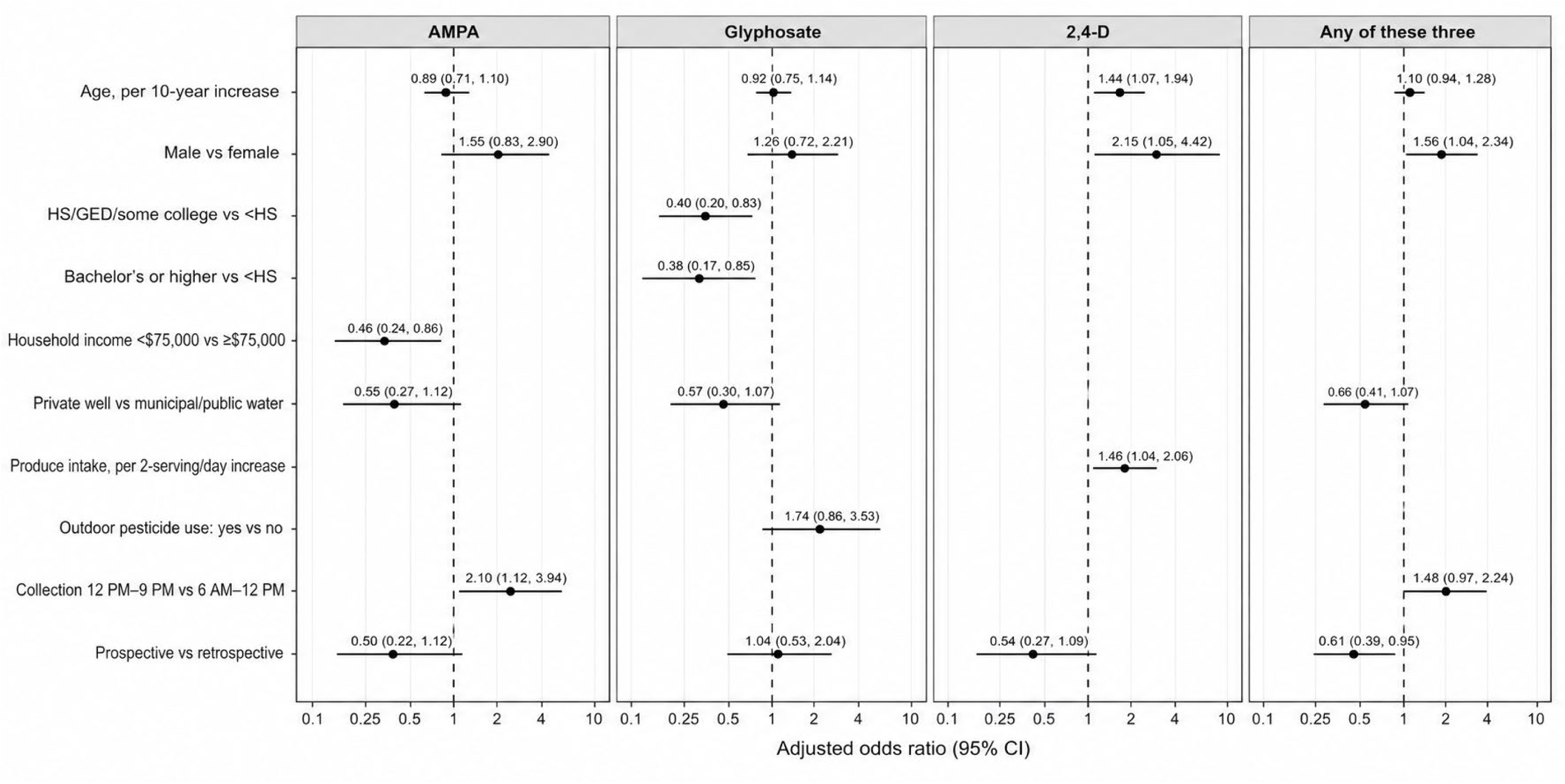
Factors associated with urinary pesticide detection in final stacked, repeated-measures models. Adjusted odds ratios and 95% confidence intervals are shown for factors retained in final generalized estimating equation models of urinary detection of AMPA, glyphosate, 2,4-D, and any of these three pesticides. Models combined retrospective (2008–2016) and prospective (2025) observations and accounted for repeated measurements within participants. All models included age, sex, and study period; additional covariates varied by outcome based on the final parsimonious model. The dashed vertical line indicates an odds ratio of 1.0; arrows indicate confidence intervals extending beyond the displayed range. AMPA, aminomethylphosphonic acid; 2,4-D, 2,4-dichlorophenoxyacetic acid; CI, confidence interval.

Age, sex, and diet were the strongest predictors of 2,4-D detection. Each 10-year increase in age was associated with 44% higher odds of detection (OR, 1.44; 95% CI, 1.07–1.94), males had approximately twice the odds of detection compared with females (OR, 2.15; 95% CI, 1.05–4.42), and each 2-serving/day increase in fruit and vegetable intake was associated with higher odds of detection (OR, 1.46; 95% CI, 1.04–2.06). Male sex was also associated with detection of any of the three most commonly detected pesticides (OR, 1.56; 95% CI, 1.04– 2.34). Detection of any of the three was less likely during prospective versus retrospective collection (OR, 0.61; 95% CI, 0.39–0.95); study period was not clearly associated with AMPA, glyphosate, or 2,4-D individually.

### Prospective-only analyses

Prospective-only analyses allowed evaluation of more detailed agricultural, dietary, and recent pesticide-contact measures that were not available retrospectively. Estimates were less precise because of the smaller sample and low number of pesticide detections, and these findings were considered exploratory (Supplementary Figure S2; Supplementary Table S8). Nevertheless, several patterns were consistent across related exposure measures and outcomes. Greater fruit and vegetable intake remained associated with AMPA detection in the final prospective model (OR, 1.40 per 2 additional eating occasions; 95% CI, 1.07–1.85). Private-well use was associated with lower odds of glyphosate detection (OR, 0.34; 95% CI, 0.12–0.97). For 2,4-D, increasing age (OR, 2.67 per 10 years; 95% CI, 1.28–5.57) and male sex (OR, 3.98; 95% CI, 1.02–15.57) were positively associated with detection.

More detailed measures of agricultural activity suggested stronger associations with working on agricultural land than simply residing near it. In univariate prospective analyses, working on field-crop agricultural land was associated with higher odds of 2,4-D detection (OR, 9.10; 95% CI, 2.13–38.84) and detection of any of the three primary pesticides (OR, 5.67; 95% CI, 1.74–18.52), whereas living on or adjacent to field-crop land was not associated with either outcome. Positive associations with field-crop work were also observed for AMPA and glyphosate, although these estimates were weaker and more imprecise. Working on specialty-crop land showed similarly positive associations with glyphosate and detection of any of the three pesticides, again with wider confidence intervals. In the final prospective model for detection of any of the three pesticides, working on field-crop agricultural land remained associated with higher odds of detection (OR, 4.71; 95% CI, 1.28–17.37), while household income <$75,000 (OR, 0.42; 95% CI, 0.19–0.94) and private-well use (OR, 0.43; 95% CI, 0.19–0.99) were inversely associated. Confidence intervals for the crop-specific agricultural measures were wide, reflecting the small number of detections and smaller exposed subgroups. Associations using the broader binary measures of working on agricultural land or living on or adjacent to agricultural land were generally in the same direction but smaller in magnitude and more precise than the crop-specific estimates.

## DISCUSSION

This study adds to the limited literature characterizing pesticide exposure in population-based samples of U.S. adults, particularly among rural residents living in agricultural regions. Using archived urine specimens collected from Wisconsin adults during 2008–2016 and repeat specimens collected approximately 9–17 years later, we examined the distribution, co-detection, temporal patterns, and determinants of urinary pesticide exposure. Overall, pesticide detection frequencies and co-detection were low, with glyphosate, its related biomarker and environmental degradation product AMPA, and 2,4-D accounting for nearly all detections. Persistence within individuals across the two collection periods was also uncommon. Among potential exposure determinants, the most consistent patterns were observed for diet and direct agricultural activity, including greater fruit and vegetable consumption and working on agricultural land, particularly field-crop land. In contrast, residential cropland density, private-well use, water filtration, and residential or household pesticide use were not consistently associated with greater pesticide detection. Together, these findings suggest that dietary and direct agricultural activities may be more important sources of pesticide exposure in this population than drinking water or residential proximity to agricultural land alone.

Detection profiles have varied substantially across recent population-based pesticide biomonitoring studies, reflecting differences in the pesticides measured, laboratory methods, analytic limits of detection, and underlying exposure patterns. Ottenbros et al. examined 37 urinary pesticide biomarkers corresponding to 27 parent pesticides among adults from the Netherlands and Switzerland and reported that detection and co-occurrence were generally low, although biomarkers of acetamiprid, chlorpropham, and flonicamid were detected in ≥40% of specimens (I. B. Ottenbros et al., 2023). In contrast, a broader study across five European countries identified 40 biomarkers corresponding to 29 pesticides, with two or more pesticides detected in 84% of samples (I. B. Ottenbros et al., 2023), and a recent study of 500 Spanish adults detected 39 of 40 evaluated biomarkers at least once, with detection frequencies ranging from <1% to 65% (López et al., 2025). The predominant biomarkers in these European studies included acetamiprid, chlorpropham, and several fungicides, differing substantially from the glyphosate, AMPA, and 2,4-D profile observed in Wisconsin. These differences likely reflect both variation in the compounds included in each analytical panel and geographic differences in pesticide use and exposure patterns.

Co-detection was also uncommon in the Wisconsin sample. The most frequent combination was glyphosate with AMPA, which is not unexpected because AMPA is a glyphosate-related metabolite and major environmental degradation product of glyphosate; AMPA can also arise from other environmental sources and therefore does not exclusively represent glyphosate exposure (Buekers et al., 2022). Ottenbros et al. similarly found relatively little co-occurrence in their adult Netherlands and Switzerland study, with the most common combination detected in only 5.5% of participants (I. B. Ottenbros et al., 2023). Importantly, the pesticide combinations differed between studies, again reflecting differences in the underlying pesticide panels and exposure environments. The low persistence observed in our paired specimens is also consistent with the short biological half-lives of many nonpersistent pesticides and with prior studies demonstrating substantial within-person variability in urinary pesticide biomarkers. In the Dutch repeated-sample component of Ottenbros et al., within-person correlations across summer and winter specimens were ≤0.3. Our specimens were separated by nearly a decade or more rather than a season, so lack of persistent detection likely reflects both the short exposure window represented by a spot urine specimen and true changes in behavior, agricultural practices, and exposure over time.

Diet was one of the more consistent potential exposure pathways identified. Greater fruit and vegetable consumption was associated with higher detection of 2,4-D in the stacked analyses and with AMPA in prospective analyses. Other recent population-based studies have similarly identified diet as an important source of nonoccupational pesticide exposure. Among HBM4EU adults, greater fruit and vegetable consumption was associated with higher urinary glyphosate in the German study population, although the association was sensitive to model specification and was not consistently observed across populations (Buekers et al., 2022). More recently, an analysis of NHANES 2015–2016 found that consumption of fruits and vegetables weighted by pesticide contamination was associated with higher urinary biomarkers of organophosphate, pyrethroid, and neonicotinoid insecticides after potatoes were excluded from the dietary exposure score (Temkin et al., 2025). Similarly, a 2026 study in rural Latvia linked consumption of specific fruits and vegetables, including apples, bananas, grapes, and pears, with increased pesticide detection (Akūlova et al., 2026a). Findings regarding organic food consumption have been less consistent. Ottenbros et al. reported lower exposure to several pesticides among adults with high consumption of organic fruits and vegetables, whereas the recent Latvian study found no significant differences between organic/homegrown and conventionally produced foods (Akūlova et al., 2026a; I. B. Ottenbros et al., 2023). We similarly did not observe a strong association with organic food consumption. However, organic consumption was measured only broadly (yes vs. no) in the retrospective survey, while more detailed frequency information was available only among the smaller prospective sample. A binary measure of organic consumption may not adequately distinguish individuals whose diets are predominantly organic from those consuming organic foods only occasionally.

The agricultural findings provide additional support for direct agricultural activity as an exposure pathway. Associations were modest when agricultural exposure was characterized broadly as living on or adjacent to agricultural land or working on agricultural land but became substantially stronger when prospective survey data were used to distinguish working on field-crop or specialty-crop land. Working on field-crop land was associated most consistently with 2,4-D and detection of any of the three predominant pesticides, with similar positive patterns for glyphosate; working on specialty-crop land also showed positive associations with several outcomes. Recent contact with vegetation or soil to which pesticides had been applied showed a similar pattern. These findings are consistent with recent occupational biomonitoring studies demonstrating substantially greater urinary pesticide exposure following direct agricultural activity. In a U.S. study, farmers with recent occupational glyphosate use had the highest urinary glyphosate concentrations, particularly those who had applied glyphosate during the previous day; application to crops, application method, and personal protective equipment use also influenced concentrations (Chang et al., 2024). More recently, a study of agricultural workers in Chile using paired urinary biomarkers and silicone wristbands identified field re-entry work as an important determinant of pesticide exposure across both matrices (DeSantiago et al., 2026). The stronger estimates observed for crop-specific work compared with broader agricultural residence or work measures in our study suggest that more detailed characterization of agricultural activities may be needed to distinguish meaningful exposure differences in general population samples.

Conversely, we found little evidence that broader residential agricultural measures or drinking-water source were major predictors of urinary pesticide detection. Percent cropland within one mile of the home was not consistently associated with detection, and living on or adjacent to agricultural land generally showed weaker associations than direct agricultural work. Ottenbros et al. similarly found no consistent associations with residential distance to agricultural or forest areas, and the broader HBM4EU mixture study found some differences associated with residence in agricultural areas but no consistent pattern across pesticides or study sites (I. B. Ottenbros et al., 2023). A 2026 study in rural Latvia provides an especially relevant comparison: although agricultural land was present within 1,000 meter of most participants’ residences, neither distance to agricultural land nor the amount of agricultural land surrounding the residence was consistently associated with urinary pesticide detection (Akūlova et al., 2026b). Private-well use was not associated with higher pesticide detection in our study and was inversely associated with glyphosate and detection of any of the three predominant pesticides in some models. Water filtration was also not consistently associated with detection. These findings do not rule out drinking-water or pesticide drift as exposure pathways for individual pesticides or highly exposed communities, but within this population they provide little evidence that either was a predominant source of exposure. Instead, the combined dietary and agricultural-work findings suggest that ingestion of pesticide residues in food and direct contact associated with agricultural activities may contribute more to the exposure patterns observed here.

Timing of urine collection is also important when interpreting these findings. Many current-use pesticides are nonpersistent and rapidly eliminated, such that a single spot urine specimen may reflect a relatively short exposure window and can be sensitive to the timing of collection (Connolly et al., 2019; LaKind et al., 2019). In our study, time of day was associated with detection, most notably for AMPA, whereas seasonal differences were less apparent. Ottenbros et al. standardized collection using first-morning voids and found no clear seasonal differences in Switzerland, while repeated summer and winter specimens in the Netherlands demonstrated substantial within-person variability (I. B. Ottenbros et al., 2023). Other U.S. biomonitoring studies of glyphosate have likewise used first-morning specimens, including recent occupational and dietary studies (Chang et al., 2024; Lucia et al., 2023). Standardizing urine collection may therefore reduce some short-term variability in future studies. The observed time-of-day association in our study underscores that differences in specimen collection may contribute to variability in detection, particularly when biomarkers have short biological half-lives.

Several strengths of this study should be considered. Few U.S. studies have characterized multiple urinary pesticides in a population-based sample with substantial representation of rural residents, and even fewer have repeat biospecimens collected from the same individuals many years apart. The availability of archived SHOW specimens made it possible to characterize pesticide exposure during 2008–2016 and repeat biomonitoring in the same participants in 2025. The study also integrated individual survey information, residential geospatial measures, and prospective questions specifically designed to characterize agricultural activities and potential pesticide exposure. The distinction between broad agricultural measures and crop-specific work and residential measures was particularly informative and provides direction for improving exposure assessment in future population-based biomonitoring studies.

However, there are several limitations. First, detection frequencies were low, limiting the number of pesticides that could be evaluated analytically and the number of predictors that could be included simultaneously in multivariable models. Low detection also limited our ability to conduct a traditional longitudinal analysis of within-person change; instead, the primary analysis used stacked, repeated-measures models to maximize available information while accounting for repeated observations. The prospective sample was smaller than the retrospective sample, and some crop-specific agricultural exposures were uncommon, resulting in large effect estimates with wide confidence intervals that should be interpreted cautiously. In addition, detailed questions regarding crop type, direct agricultural activities, recent contact with pesticide-treated vegetation or soil, and recent dietary intake were collected only prospectively. Retrospective SHOW surveys were designed as general health surveys rather than pesticide-specific exposure assessments and therefore contained less detailed information regarding occupational agricultural activities and recent diet. The older age distribution of the prospective sample may also have reduced representation of adults currently engaged in agricultural work and, consequently, current occupational pesticide exposures. Prospective follow-up was also conducted within a five-month field period, and 23.7% of invited participants completed a study visit. Limited scheduling opportunities and differential participation may therefore have reduced representativeness of the prospective sample. Second, spot urine collection was not standardized to a first-morning void. Collection time was associated with some pesticide outcomes, suggesting that specimen timing contributed to exposure variability. Retrospective specimens were collected throughout the year, whereas nearly all prospective specimens were collected during the growing and harvest season, limiting direct assessment of seasonality at follow-up. Finally, although household indoor and outdoor pesticide use and frequency could be harmonized across study periods, these measures did not identify strong associations with urinary detection and may not have captured the specific chemical products, amount applied, or timing of use relative to urine collection.

In conclusion, pesticide detections were generally uncommon in this population-based sample of Wisconsin adults, with glyphosate, AMPA, and 2,4-D accounting for most detected exposures and little evidence of persistent detection within individuals over time. Dietary intake and direct agricultural work, particularly work involving specific crop types, were the clearest potential exposure pathways, whereas residential cropland, private-well use, and household pesticide use were less consistently associated with detection. These findings demonstrate both the value and the challenges of pesticide biomonitoring in general population studies and support future studies using larger samples, standardized repeated urine collection, and more detailed measures of recent diet, crop-specific agricultural activities, and pesticide use to better characterize sources and temporal variability of exposure.

## Supplementary Materials

Supplementary Materials can be viewed online here: https://uwmadison.box.com/v/Wisconsin-Rural-Health-Study

## Data Availability

Limited data produced as part of this present study are available upon reasonable request to the authors. Baseline data and specimen used from the SHOW cohort are available through the UW-Madison Real-World Evidence for Advancing Community Health (REACH) program: https://reach.med.wisc.edu/

## Funding

This study was funded by the Centers for Disease Control (CDC), award NU88EH001346. Authors or their institutions did not receive payment or services from a third party for any aspect of the submitted work.

## Conflict of Interest

The authors have declared no conflict of interest.

## Disclosures

The authors have no disclosures.

## Acknowledgements

The authors would like to thank the SHOW participants for completing the study. The authors would also like to thank the UW REACH program for continuing to maintain the SHOW cohort and provide their research study services.

## Notes

### Competing Interest Statement

The authors have declared no competing interest.

### Author Declarations

The Health Sciences Institutional Review Board of the University of Wisconsin-Madison gave ethical approval for this work.

